# COVID-19 vaccine uptake, predictors of vaccination, and self-reported barriers to vaccination among primary school teachers in Poland

**DOI:** 10.1101/2021.07.11.21260317

**Authors:** Marta Malesza, Karolina Sobolewska

## Abstract

It has been proposed that teachers, like healthcare workers, constitute a strategic target for COVID-19 vaccine programs as immunization is a key element in protecting both them and their pupils. The present study examined vaccine uptake among primary-school teachers and sought to identify factors associated with it. A sample was recruited from 553 Polish primary schools, and data were collected at two time points: December 2020 and March 2021. Associations between vaccine uptake among teachers and their attitudes toward COVID-19 vaccination were assessed through multivariate logistic regression. 6152 participants completed both baseline and follow-up surveys. Of these, 4502 (73.2%) reported their intention at baseline to receive a COVID-19 vaccination, if available; at follow-up, 3894 (86.5%) of the same 4502 reported having received the vaccination. A significant association was revealed between vaccine uptake and perceived severity, self-efficacy, and social norms. The principal driver for vaccine acceptance was the wish to avoid contracting the disease. Conversely, the principal driver for vaccine refusal was concern about side effects and safety. A strong association exists between intention to receive the COVID-19 vaccine and actual uptake. Future COVID-19 immunization programs may benefit from a stronger understanding of the factors associated with vaccine uptake among this cohort.

## Introduction

Coronavirus Disease-19 (COVID-19) was first identified in December 2019 in the Chinese city of Wuhan, thereafter spreading rapidly around the globe. The World Health Organization (WHO) declared a Public Health Emergency of International Concern on 30 January 2020 [1] and a pandemic on March 11. In Poland, of the 1,330,543 people who contracted the virus, 29,502 had died as of 5 January 2021. The Polish pandemic preparedness plan is based on WHO recommendations and depends largely on a vaccination program, under which mass vaccinations of teachers began toward the end of January 2021. A government official has said that all willing teachers in Poland should be vaccinated against the coronavirus by the end of March 2021 [2].

As seen in seasonal influenza pandemics, virus transmission among school-aged children has been key to the spread of COVID-19 [3]. The 15,000 public and private Polish primary schools together cater to more than 3 million pupils and employ staff totalling approximately 223,000. Given the size of the school population, the necessarily close contact among individuals, and the difficulty in enforcing boundaries, there is considerable concern that schools will see intense, centrifugal outbreaks [4]. Despite this concern, little data has been gathered regarding the association in adult individuals, including educators, between intention to be vaccinated and real-life uptake of the vaccine. A handful of studies have sought to shed light on factors driving willingness to be vaccinated against COVID-19 under the condition that the vaccine is freely available to everyone [5-9]. However, most were either carried out at pre-pandemic stage, before the vaccine became available, or have only evaluated specific cohorts, such as healthcare workers. Hence, the existing research in this area may not fully explain the latest uptake rates, given possible changes in public perceptions of both pandemic and vaccine during the course of the outbreak [5-7].

Similarly, limited data has been collected concerning vaccine attitudes and uptake among teachers, although a body of US research was undertaken in the 2000s. One 2009 survey of teachers in the state of Georgia reported a rate of influenza immunization of nearly 53% [10], while a similar study by Gargano et al. found 78% of respondents had been vaccinated against seasonal influenza in the 2008/2009 season [11]. Data collected in Ohio for all types of staff employed at educational institutions indicated a 58% vaccination rate for the 2012/2013 season [10]. A study conducted by Vaughn et al. found that 36% of teachers surveyed had been vaccinated five times over five years, which is comparable with the rate seen in the general adult population of the US [12].

Multiple researchers have examined predictors of agreement to receive a vaccination against seasonal influenza among healthcare workers. This body of research may be of value to the present study because of the similarity in working environment of the two cohorts: Members of both professions work in settings where disease spreads easily and hence are prone to contracting and transmitting infections. The most common drivers for intent to receive the seasonal influenza vaccine found in studies of healthcare workers were the desire to protect self, family, or patients [13-15]. Teachers, like healthcare workers, may be driven by the same urge to protect family, friends, and/or students against infection.

A separate but related body of work has examined predictors of intent to receive the H1N1 influenza vaccine offered to adults in 2009, finding that the principal drivers of acceptance were perceived risk, perceived safety, and having been vaccinated against seasonal influenza in the past [16-18]. Healthcare workers surveyed about their intent to receive the 2009 H1N1 influenza vaccine similarly cited safety and efficacy concerns as drivers of acceptance [19-20]. The COVID-19-related literature expands on these findings by suggesting that greater compliance with vaccine programs will follow from vaccine recipients perceiving the illness to have severe consequences or if they already have a severe chronic illness [5-9].

No matter how effective a vaccine is, it will have little impact if there is low take-up. If effective public health pandemic strategies are to be developed, it is vital to understand the different factors which motivate vaccination compliance. However, to date no research has been conducted to evaluate uptake of the COVID-19 vaccine among teachers, even though this cohort may be among the groups targeted by immunization programs. The current study aims to investigate drivers motivating or preventing uptake of the COVID-19 vaccine among Polish primary-school teachers. The study sought in particular to evaluate the association between baseline intention to be vaccinated against COVID-19 and factors driving later compliance or non-compliance with the COVID-19 vaccination program.

## Methods

### Study population and sampling

A sample of Polish primary-school teachers was recruited to participate in this study. Data collection was carried out by emailing surveys to respondents at two time points. The following eligibility criteria were applied: (1) being employed as a teacher in a participating primary school; and (2) provision of online informed consent to participate. The first survey round was administered in December 2020, and the second in March 2021. The sample was nationally representative of this cohort, as per the guidelines contained in the Declaration of Helsinki. The Institutional Review Board of University of Economics and Social Sciences in Warsaw, Poland approved all procedures involving human subjects.

Informed consent to participate in this study was given by all respondents. Respondents were recruited via random quota sampling of primary schools in order to achieve a study population which represented schools from each of the Polish regions. Poland’s six principal geographic regions encompass 16 administrative units (*voivodeships*), divided along on historic, cultural, economic, and geographic lines, each of which is subdivided into counties. A stratified sampling method was employed, and invitations to participate were sent to randomly-chosen primary schools. There were two levels of sampling: first, sampling of counties within voivodeships, whereby five counties from each voivodeships (total=80 counties) were chosen; and second, sampling of primary schools within counties, whereby 10 schools for each of the counties were chosen (total=800 schools). The principal of each of the 800 resulting primary schools was asked if their school would be willing to participate on a voluntary basis. Those willing to do so then informed their staff of the invitation, making it known that participation was voluntary. Thereafter, an electronic link to the dedicated questionnaire was sent. The questionnaire was in anonymized form; hence, no data could be collected from which respondents could be identified and no items addressed issues potentially seen as personal or sensitive. Respondents were not offered compensation, monetary or otherwise, for their participation. Of the 800 schools contacted, 553 (69.1%) were willing to participate.

Completion rate for the baseline survey was 76.6% (7890/10295), and completion rate for the follow-up survey was 80.0% 6152/7890). Of the total 6152 participants considered, that is, those who completed both surveys, 79.3% were female and 20.7% were male. They ranged in age between 23 and 59 (*M*=39.92 and *SD*=13.45) and had taught for between 1 and 32 years (*M*=12.75 and *SD*=8.51). A little over half the sample (52.6%; n=3238) had less than 10 years’ teaching experience, and the rest (47.4%; n=2914) had up to 32 years’ such experience. In terms of marital status, 3522 (57.3%) were married or cohabiting, 1897 (30.8%) were single, 649 (10.5%) were divorced, and 84 (1.4%) were widowed. As concerns the grades taught, 2064 (33.5%) taught lower primary (Grades Preparatory, 1, 2, and 3), 2107 (34.3%) taught middle primary (Grades 4 and 5), 1640 (26.7%) taught upper primary (Grades 6, 7 and 8), and the remaining 341 (5.5%) taught specialist classes to several grades.

## Materials

Similar survey materials were sent to respondents at baseline and follow-up. Data were gathered via a self-administered, online questionnaire, sent to respondents at their school email addresses. Survey items were designed to evaluate demographic, behavioral, and psychosocial factors driving acceptance or refusal of the COVID-19 vaccination. The Health Belief Model (HBM) [21] and Integrated Behavioral Model (IBM) informed the design of the psychosocial survey items [22]. Items were adapted from relevant existing surveys [23-24].

### Vaccination uptake

The baseline survey consisted of one item asking whether respondents would agree to be vaccinated once a vaccine against COVID-19 became available (“Would you consider getting vaccinated against the novel coronavirus once the vaccine becomes available?”) There were two possible answers: yes or no. The follow-up survey assessed vaccine uptake through the item “Have you already been vaccinated against COVID-19?” Respondents answering “no” were then asked “Have you already registered for the vaccine and are now waiting your turn?” to evaluate their attitudes towards the vaccination.

### Psychosocial variables

Respondents’ attitudes toward and beliefs regarding the COVID-19 vaccination were evaluated through seven psychosocial variables. (See Appendix 1 for a detailed description of items, scale ranges, and Cronbach’s alphas.) Respondents were asked to answer items addressing psychosocial constructs using 5-point Likert scales where 1=strongly disagree and 5=strongly agree. Responses for each item were then summed to give the range for each construct.

Six psychosocial measures drew on the guidelines provided by the HBM. The first two addressed respondents’ perceptions of the severity of, and their vulnerability to, COVID-19 infection. The next two examined perceptions of benefits of, and barriers to, having the vaccination, and the last two concerned perceptions of self-efficacy and cue to action in receiving the vaccine. IBM theory was drawn on to guide the final variable of social norms.

### Reasons for vaccine acceptance or refusal

Respondents who indicated they had received the COVID-19 vaccination were shown a list of possible reasons for having done so and asked to answer “yes” or “no” to indicate whether each had been a driver in their decision. Respondents who had not received the COVID-19 vaccination were shown a list of possible reasons for refusal and asked to answer “yes” or “no” to indicate whether each was a driver in their case. The possible responses were not mutually exclusive. Table 2 presents the evaluated reasons for compliance with and refusal of vaccination.

### Demographic factors

Respondents were also asked for demographic information, including gender, age, marital status, grade taught/primary job at their place of employment, and whether or not they had children. Two questions were designed to assess respondents’ health status: “How good is your health generally?” (very good/good/bad/very bad); and “Do you have any of the following chronic illnesses?” (cancer, heart disease, lung disease, liver or kidney disease, and any other illness).

### Data analysis

The following steps were taken to analyze the data gathered. First, questions related to psychosocial constructs were combined into scales, for each of which Cronbach’s alpha was calculated to evaluate internal consistency, as shown in Appendix 1. Thereafter, demographic, behavioral, and psychosocial variables concerning COVID-19 vaccination were assessed by means of univariate descriptive analyses. In a third step, chi-square analyses were undertaken to evaluate the association between baseline intention to be vaccinated against COVID-19 and self-reported uptake at follow-up. Next, bivariate analyses were used to evaluate the demographic and psychosocial correlates of uptake. Significant variables (p < 0.05) in the bivariate analyses were entered into multivariate logistic regression analyses to evaluate correlates of vaccine uptake. Lastly, a frequency analysis evaluated drivers of vaccination compliance among those respondents who had indicated they had been vaccinated at follow-up and vaccination non-compliance among those who indicated they had not been vaccinated at follow-up.

## Results

### Baseline vaccination intention and uptake

Of the 6152 respondents considered (i.e., those who completed surveys at both time points), 4502 (73.2%) signalled their intent to be vaccinated against COVID-19 at baseline, if a vaccine were available. Of these 4502, 3894 (86.5%) indicated they had actually received the vaccination at follow-up (Chi-square = 18.9, p < 0.001).

### Correlates of COVID-19 vaccination

Table 1 shows the correlates of COVID-19 vaccination among teachers. Bivariate analyses reveal that correlates of COVID-19 vaccine uptake include the following: *perceived severity* of COVID-19 infection; *perceived benefits of* and *barriers to* COVID-19 vaccination; *self-efficacy* for COVID-19 vaccination; and *social norms* concerning COVID-19 vaccination. As demographic variables, *cue to action* and *perceived susceptibility* to COVID-19 infection were found to be not significant, they were not considered in the multivariate analyses. Out of five variables four were significant in the multivariate analyses – they showed a significant association with receipt of the vaccination. Individuals who *perceived greater severity* of COVID-19 infection were more likely to receive vaccination. Respondents who *perceived more barriers* to getting a COVID-19 vaccination were also less likely to receive one. Respondents with more positive *social norms* in regard to the vaccination and greater *self-efficacy* were more likely to receive one.

**Table 1.**
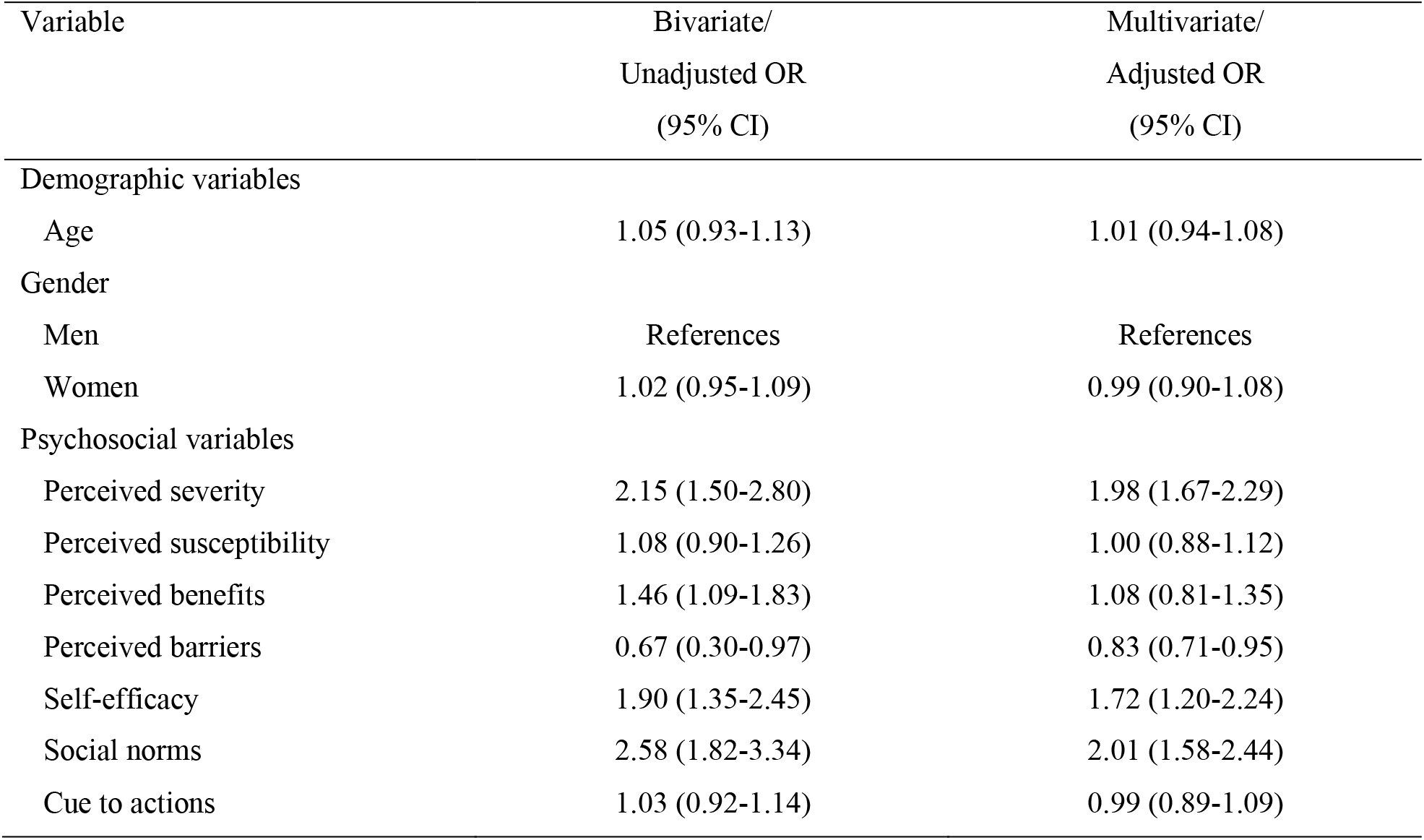
Correlates of COVID-19 vaccine uptake among primary-school teachers

| Variable | Bivariate/<br>Unadjusted OR<br>(95% CI) | Multivariate/<br>Adjusted OR<br>(95% CI) |
| --- | --- | --- |
| Demographic variables |  |  |
| Age | 1.05 (0.93-1.13) | 1.01 (0.94-1.08) |
| Gender |  |  |
| Men | References | References |
| Women | 1.02 (0.95-1.09) | 0.99 (0.90-1.08) |
| Psychosocial variables |  |  |
| Perceived severity | 2.15 (1.50-2.80) | 1.98 (1.67-2.29) |
| Perceived susceptibility | 1.08 (0.90-1.26) | 1.00 (0.88-1.12) |
| Perceived benefits | 1.46 (1.09-1.83) | 1.08 (0.81-1.35) |
| Perceived barriers | 0.67 (0.30-0.97) | 0.83 (0.71-0.95) |
| Self-efficacy | 1.90 (1.35-2.45) | 1.72 (1.20-2.24) |
| Social norms | 2.58 (1.82-3.34) | 2.01 (1.58-2.44) |
| Cue to actions | 1.03 (0.92-1.14) | 0.99 (0.89-1.09) |

### Reasons for vaccination and non-vaccination against COVID-19

Table 2 summarizes the drivers for compliance and non-compliance with the COVID-19 immunization program among respondents. The four reasons most commonly cited for compliance by those participants who had been vaccinated (n=3894) were the desire to avoid being infected by COVID-19; the wish to protect family members against the disease; knowing someone who got sick from the COVID-19; and having heard news reports about the pandemic. Among respondents who did not get vaccinated (n=2258), the main reasons were concerns about vaccine safety and fear of vaccine side effects (answered by 97.9% and 97.6% respondents, respectively). Also 74.1% of individuals said they do not want to be used as the subject of an experiment (do not want ‘‘to be a guinea pig’’). 43.4% claimed that they do not need vaccine since they are not in a risk group. Almost one-third of respondents (37.9%) stated that COVID-19 is not a severe disease. Almost the same percent of people (35.6%) stated that vaccines are ineffective.

**Table 2.** Reasons for acceptance or refusal of COVID-19 vaccine

| **Reasons for acceptance (N = 3894)** | **N (%)** |
| --- | --- |
| Protecting myself to avoid sickness | 3774 (96.9) |
| Protecting my close relatives | 3011 (77.4) |
| Know someone who got sick from the COVID-19 | 2520 (64.7) |
| Heard about it on the news | 2243 (57.6) |
| The vaccine will stop the outbreak | 1908 (49.0) |
| Vaccines are safe | 1573 (40.4) |
| Vaccines have no side effects | 1570 (40.3) |
| Getting vaccinated is convenient and quick | 1272 (32.7) |
| Protecting myself to avoid work absenteeism | 1249 (32.1) |
| Vaccination is recommended by public authorities/ health officials | 1205 (30.9) |
| Getting vaccinated is a civic duty | 87 (2.2) |
| **Reasons for refusal (N = 2258)** |  |
| Vaccines are not safe enough | 2210 (97.9) |
| Vaccines have side effects | 2203 (97.6) |
| I don't want to be a guinea pig | 1673 (74.1) |
| I don't need it, since I am not in a risk group | 980 (43.4) |
| COVID-19 is not a severe disease | 856 (37.9) |
| Vaccines are ineffective | 803 (35.6) |
| Did not know I was eligible for the COVID-19 vaccine | 675 (30.0) |
| I dislike the shots | 230 (10.2) |
| Getting vaccinated is inconvenient and too long | 207 (9.2) |
| I have medical reasons to avoid vaccines | 184 (8.1) |
| I don't know | 46 (2.0) |

## Discussion

Gaining a deeper understanding of how teachers perceive the COVID-19 vaccine may enhance response planning and management strategies both during and after a pandemic. Consequently, the present study has sought to examine which factors have impacted vaccine compliance and non-compliance during the COVID-19 pandemic among teachers at Polish primary schools, as well as to assess real-life vaccine uptake. The two specific goals of this research were to measure intention to receive the vaccine against self-reported uptake of it; and to investigate why respondents in this cohort agreed or refused to be vaccinated against COVID-19.

Data gathered reveals that 63.3% of respondents reported having been vaccinated. This study offers a contribution to the related literature by investigating both intent to receive a COVID-19 vaccine and actual vaccine uptake, as well as drivers of vaccine compliance. Of the respondents who completed the surveys at both timepoints, 86.5% of those who stated their intention to be vaccinated did, in fact, receive the vaccination at a later date, indicating a robust association between intention to be vaccinated and actual vaccination uptake during the COVID-19 pandemic. It is hoped that these results will inform future studies on interventions aimed at adults who have stated their intention to be vaccinated but may fail to put this intention into action. Besides, more remarkable, there is a significant gap between intention and actual behavior to get vaccinated. A recent investigation showed that the willingness for influenza vaccination was 45% in general population [25], while the actual vaccination coverage was 9.4%, which was reported by a meta-analysis [26]. These indicate that there are barriers existed from intention to behavior besides the cognitive factors, which affected the vaccination willingness, such as not receiving recommendation from doctor and not having cost-free vaccination [25]. Previous studies have examined factors associated with intention of COVID-19 vaccination [5-9]. However, less is known about COVID-19 vaccination intention, actual uptake and the related factors. Thus, the present study is also the first to focus on this cohort (primary-school teachers), despite the fact that they constitute a population of considerable importance for immunization programs [2-3]. Vaccinating this cohort against COVID-19 is a key means of extending protection against the virus not only to them but also to their pupils.

A commonality between educators and healthcare professionals is that both occupy key spaces in their communities. Both groups, moreover, work in conditions where disease can spread rapidly, exposing teachers to higher risk for infection and, hence, of passing the virus on to others. Recent research indicates that over 70% of health workers have received the COVID-19 vaccine [9; but see 27 for contrary results], while the present study reveals a similar uptake among teachers. Significant predictors for COVID-19 vaccination were revealed to be perceived severity and self-efficacy. Compliance with public health measures is largely dependent on individual perceptions of threat [28]. The focus of the present study was vaccine uptake; however, our results align with studies assessing intent to receive a COVID-19 vaccine among healthcare professionals, which found the same association between vaccine non-compliance and lack of perceived personal need or an individual’s belief that they were not at risk [9]. It is suggested that lack of perceived severity should be addressed in information campaigns, which should demonstrate the strong potential that teachers will transmit infection to their pupils. Such campaigns should also forcefully address the other aims of the COVID-19 immunization program, including the protection of other members of the community by means of herd immunity.

Multivariate analyses also revealed that perceived barriers and social norms regarding vaccination were correlated with COVID-19 vaccine uptake. Among perceived barriers, respondents indicated beliefs that the vaccine would make recipients sick, and that it was ineffective in preventing COVID-19. Findings of a survey of healthcare workers asked about their perceptions of and attitudes to the COVID-19 vaccine revealed similar beliefs [9]. In order to improve vaccine uptake, it is necessary that educational campaigns are run during periods when anxiety rates and risk perceptions are low concerning the vital need for infection control. Moreover, disseminating information about the effectiveness of the COVID-19 vaccine may lead to greater active uptake, rather than mere intention or acceptance. A significant association was also revealed between vaccine receipt and social norms, as respondents indicated their belief that having the vaccine would win them approval from medical providers, family members, and others. These findings, too, align with those of other research which found that social norms were a predictor of intention to be vaccinated as well as actual vaccination receipt [29-32]. Given the key role played by normative beliefs in the cohort studied here, campaigns urging take-up of the COVID-19 vaccination may benefit from showing endorsements of the vaccination from trusted individuals such as physicians and leveraging acceptance of this social norm. One interesting finding is that perceived susceptibility did not significantly predict uptake of the COVID-19 vaccine, despite the close contact between the surveyed population and children, who are known to be key transmitters of the disease.

The most important drivers of vaccine acceptance were the desire not to become sick by contracting COVID-19 and hearing news reports about the pandemic. The latter reason underlines the need for effective public health communications, as public perceptions of both the vaccine and the severity of COVID-19 have been substantially influenced by media reporting [5-9]. Hence, the power of the media should be harnessed very early on to encourage vaccine take-up. Principal drivers for non-vaccination against COVID-19 were safety concerns, a perception that the vaccine was unnecessary, apprehension that it was not safe, and fears to do with its newness. These findings align with other studies of acceptance of the COVID-19 vaccine [5, 8] and indicate a need to persuade the public that a vaccine is both safe and effective. Many people, moreover, have limited understanding of how the new vaccine was developed. If infectious diseases are to be effectively controlled, widespread acceptance of new vaccines is vital.

Like healthcare professionals, teachers provide a vital community service and should be strategically targeted by COVID-19 immunization programs. Educators are at greater risk of contracting and transmitting the disease, placing themselves, their families, and their pupils at risk. To our knowledge, this is the first study to evaluate drivers of seasonal and COVID-19 vaccination uptake among teachers. When pandemic vaccines are promoted to the general public, psychosocial factors, particularly perceived barriers, should be borne in mind. In the case of COVID-19 vaccination programs, a greater focus on safety, risk, and social norms may lead to higher uptake. The data presented in this paper may inform educational programs designed to boost uptake of the COVID-19 vaccine and also be of value to post-pandemic planning.

## Limitations

Our work suffers from limitations. First, the cross-sectional survey design necessarily represents a snapshot in time, rather than the evolving landscape of the teachers’ attitudes about COVID-19 vaccination. All the information obtained was self-reported and reporting bias always exists. Although the data was collected from the heterogenous group, we targeted individuals who are willing to participate and give their answers. The individual’s opinion also can be unstable. Any unexpected event could lead to drastic change in their opinion about the vaccination. Next, even though the questionnaire was anonymous, it is still possible that a social-desirability bias tainted respondents’ answers to the questionnaire about intentions and behaviors. The final limitation concerns the timing of the survey that might have led to both an overestimate of willingness to receive the vaccination and an underestimate of the vaccine coverage rate among Polish teachers population since the controversy about the efficacy, safety, and necessity of the vaccine against COVID-19.

## Data Availability

The data that support the findings of this study are available from the corresponding author upon reasonable request.

## Declarations

### Institutional Review Board Statement

The project was approved by the local ethics committee of the University of Economics and Social Sciences in Warsaw, Poland.

---

**Appendix 1.**
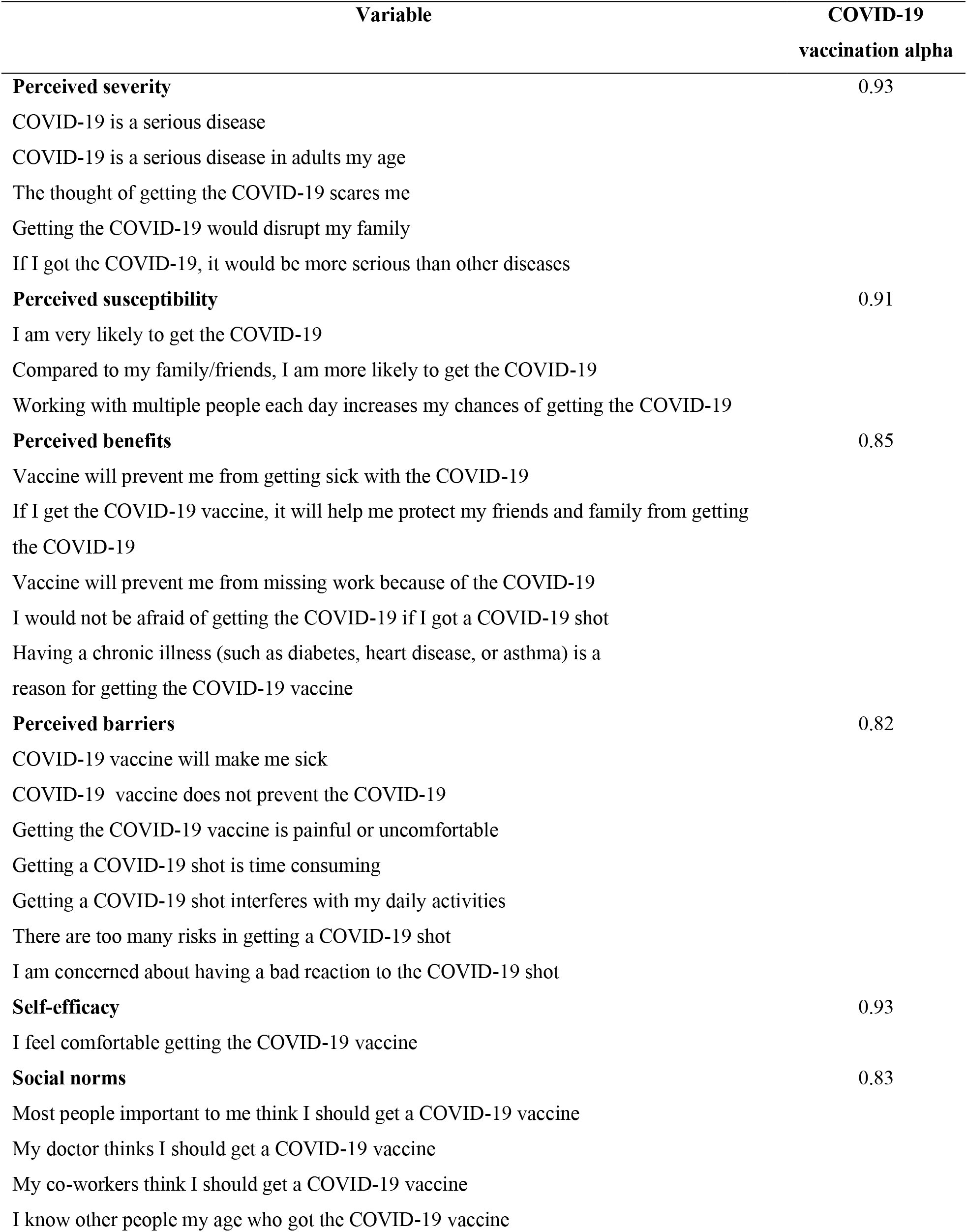
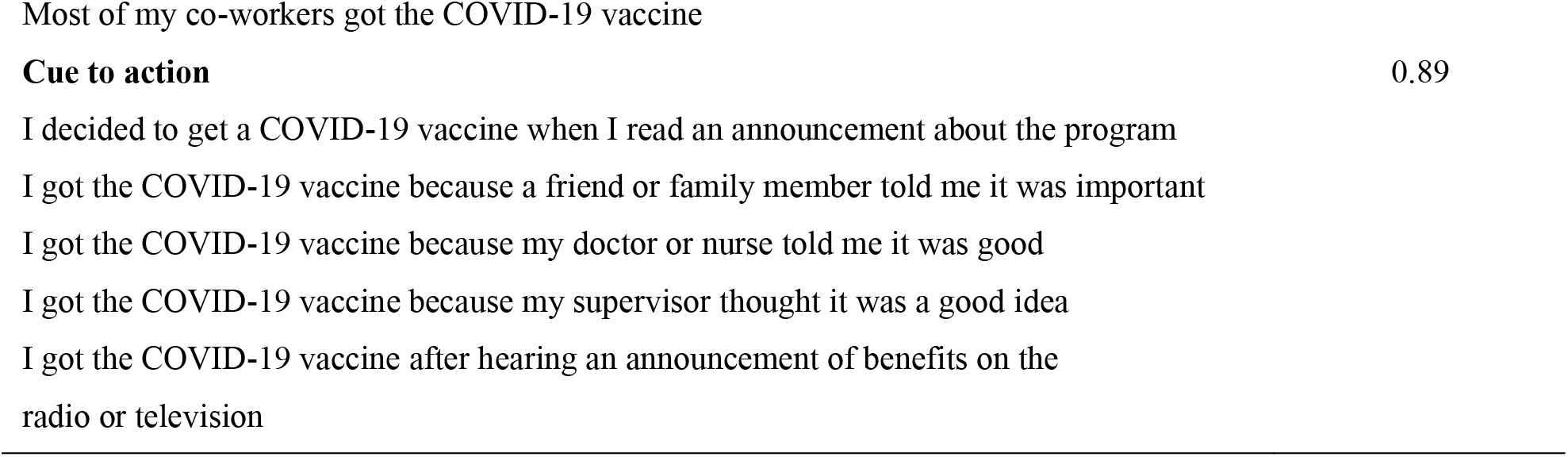
Survey items for psychosocial variables

| Variable | COVID-19<br>vaccination alpha |
| --- | --- |
| **Perceived severity** | 0.93 |
| COVID-19 is a serious disease |  |
| COVID-19 is a serious disease in adults my age |  |
| The thought of getting the COVID-19 scares me |  |
| Getting the COVID-19 would disrupt my family |  |
| If I got the COVID-19, it would be more serious than other diseases |  |
| **Perceived susceptibility** | 0.91 |
| I am very likely to get the COVID-19 |  |
| Compared to my family/friends, I am more likely to get the COVID-19 |  |
| Working with multiple people each day increases my chances of getting the COVID-19 |  |
| **Perceived benefits** | 0.85 |
| Vaccine will prevent me from getting sick with the COVID-19 |  |
| If I get the COVID-19 vaccine, it will help me protect my friends and family from getting the COVID-19 |  |
| Vaccine will prevent me from missing work because of the COVID-19 |  |
| I would not be afraid of getting the COVID-19 if I got a COVID-19 shot |  |
| Having a chronic illness (such as diabetes, heart disease, or asthma) is a reason for getting the COVID-19 vaccine |  |
| **Perceived barriers** | 0.82 |
| COVID-19 vaccine will make me sick |  |
| COVID-19 vaccine does not prevent the COVID-19 |  |
| Getting the COVID-19 vaccine is painful or uncomfortable |  |
| Getting a COVID-19 shot is time consuming |  |
| Getting a COVID-19 shot interferes with my daily activities |  |
| There are too many risks in getting a COVID-19 shot |  |
| I am concerned about having a bad reaction to the COVID-19 shot |  |
| **Self-efficacy** | 0.93 |
| I feel comfortable getting the COVID-19 vaccine |  |
| **Social norms** | 0.83 |
| Most people important to me think I should get a COVID-19 vaccine |  |
| My doctor thinks I should get a COVID-19 vaccine |  |
| My co-workers think I should get a COVID-19 vaccine |  |
| I know other people my age who got the COVID-19 vaccine |  |

